# Walking pace optimizes conventional cardiovascular disease risk prediction models among vulnerable subpopulations: a prospective cohort study

**DOI:** 10.1101/2025.03.13.25323929

**Authors:** Li Wang, Xueqin Li, Xueqing Jia, Yuyang Zheng, Jing Shao, Zuyun Liu

## Abstract

**Background:** Grip strength and walking pace are increasingly recognized as potential indicators of adverse health outcomes for general population. However, their clinical utility in the primary prevention of cardiovascular disease (CVD) amoung various subpopulations remains uncertain. This study aimed to evaluate performances of four conventional CVD prediction models across subpopulations with varying grip strength or walking pace and determine added predictive value of grip strength and walking pace.

**Methods:** A total of 206,371 individuals without CVD (aged 40-69 years) from UK Biobank (UKB) were included. Four conventional CVD prediction models (Framingham, Reynolds, ASSIGN, and Pooled Cohort Equations) were used to estimate 10-year CVD risk. Model performance was compared across diverse subpopulations defined by age, grip strength, or walking pace using Harrell’s concordance index (C index) and calibration plot in UKB. Added predictive value of grip strength and walking pace was evaluated using C index change and net reclassification improvement (NRI) in UKB and further validated in English Longitudinal Study of Ageing (ELSA).

**Results:** 19,664 cases of incident CVD were registered during follow-up period (mean 12.85 years [standard deviation 2.74]). All four models performed inferior among vulnerable subpopulations characterized by advanced age, low grip strength or slow walking pace. C index for ASSIGN model was 0.659 (95%CI, 0.646-0.672) in low grip strength subpopulation vs 0.702 (95%CI, 0.699-0.706) in normal grip strength subpopulation, 0.646 (95%CI, 0.635-0.657) in slow walking pace subpopulation vs 0.701 (95%CI, 0.698-0.705) in normal walking pace subpopulation, and 0.624 (95%CI, 0.614-0.634) in old subpopulation vs 0.701 (95%CI, 0.689-0.712) in young subpopulation. Adding walking pace to ASSIGN model improved its discriminative ability in vulnerable subpopulations, i.e., those with low grip strength (NRI, 0.046 [95%CI, 0.009-0.100]) and old subpopulation (NRI, 0.035 [95%CI, 0.020-0.048]). However, grip strength did not show significant added predictive value for CVD in these subpopulations. The finding was similar in ELSA.

**Conclusions:** Conventional CVD prediction models underperformed in vulnerable subpopulations; Walking pace, but not grip strength, conferred substantial incremental information to these models, paricularly those with advanced age or low grip strength. This current finding offers novel insights into the role of physical function in the primary prevention of CVD.

**Clinical Perspective:** *What Is New?:*

- Conventional CVD risk models did not work well in vulnerable subpopulations characterized by advanced age, low grip strength or slow walking pace.
- Walking pace, but not grip strength, conferred substantial incremental information to the established CVD risk models in vulnerable subpopulations, especially those with advanced age or low grip strength.

*What Are the Clinical Implications?:*

- Walking pace, a simple, easy-to-implement and low-cost measures, has the potential to better predict CVD risk in vulnerable subpopulations, supporting more accurate identification of high-risk population, personalization of care plans, and equitable health policies.

## Introduction

Cardiovascular diseases (CVD) are the leading cause of death globally, responsible for an estimated 19.8 million deaths in 2022[1], increasing the global healthcare burden. Early identification of individuals at high risk for CVD shapes primary preventive strategies to reduce the risk of incident CVD[2]. Risk scores based on conventional risk factors for CVD, e.g., Framingham risk profile[3], Reynolds risk score[4], ASSIGN score[5], and Pooled Cohort Equations (PCEs)[6], are widely available to estimate an individual’s risk for future CVD events. However, these conventional CVD models are established based on the general population and are found to be inappropriate in vulnerable subpopulation[7–9], such as the older adults. Compared with younger adults, older adults have a higher risk of CVD and a combined 13-fold greater risk of ischemic heart disease (IHD), ischemic stroke (IS), and peripheral artery disease (PAD) based on the 2019 Global Burden of Disease Study[10]. Therefore, revisiting conventional CVD risk prediction models and improving their predictive accuracy in vulnerable subpopulations are the priorities of cardiovascular nursing and public health.

Physical fuction is a remarkable indicator to measure health condition, especially in older adults[11–13]. Grip strength and walking pace are easily available measurements of physical function. Several studies have reported strong associations of grip strength with incident CVD events, as well as walking pace with all-cause mortality in older patients with CVD[14, 15]. However, there is little data on performances of these conventional CVD risk prediction models in vulnerable subpopulations, particularly defined by low grip strength or slow walking pace. Also, whether adding grip strength or walking pace would improve their predictive value in vulnerable subpopulations remains unclear.

In the current study, we aimed to evaluate the performance of four conventional CVD prediction models (Framingham, Reynolds, ASSIGN, and PCEs) with particular focus on vulnerable subpopulations defined by low grip strength and slow walking pace. We also investigated the potential improvement in model performance by adding grip strength or walking pace.

## Methods

### Study Design and Population

The UK Biobank (UKB) is a population-based cohort comprising more than 500,000 individuals 40 to 69 years of age, whose design has previously been described[16]. The recruitment was conducted between 2006 and 2010, where participants were invited to attend 1 of the 22 centers located throughout England, Scotland, and Wales for baseline assessments. Information was collected via touch-screen questionnaires, biological samples, physical measurements, and linked medical or death register records. Written informed consent was provided by all participants, and the study had received ethical approval from Northwest-Multicentre Research Ethical Committee. From the initial sample of 502,185 participants, those with prevalent CVD (defined as heart failure, atrial fibrillation, ischemic heart disease, and stroke cases diagnosed prior to the date of baseline assessment) were excluded (n=34,398). Participants without available records of grip strength, walking pace, or other variables were also excluded (n=261,416). In total, 206,371 individuals who were free from CVD at baseline and had complete information on all variables were eligible for the analysis. The details of the selection process are shown in **Figure 1 (A)**.

**Figure 1.**
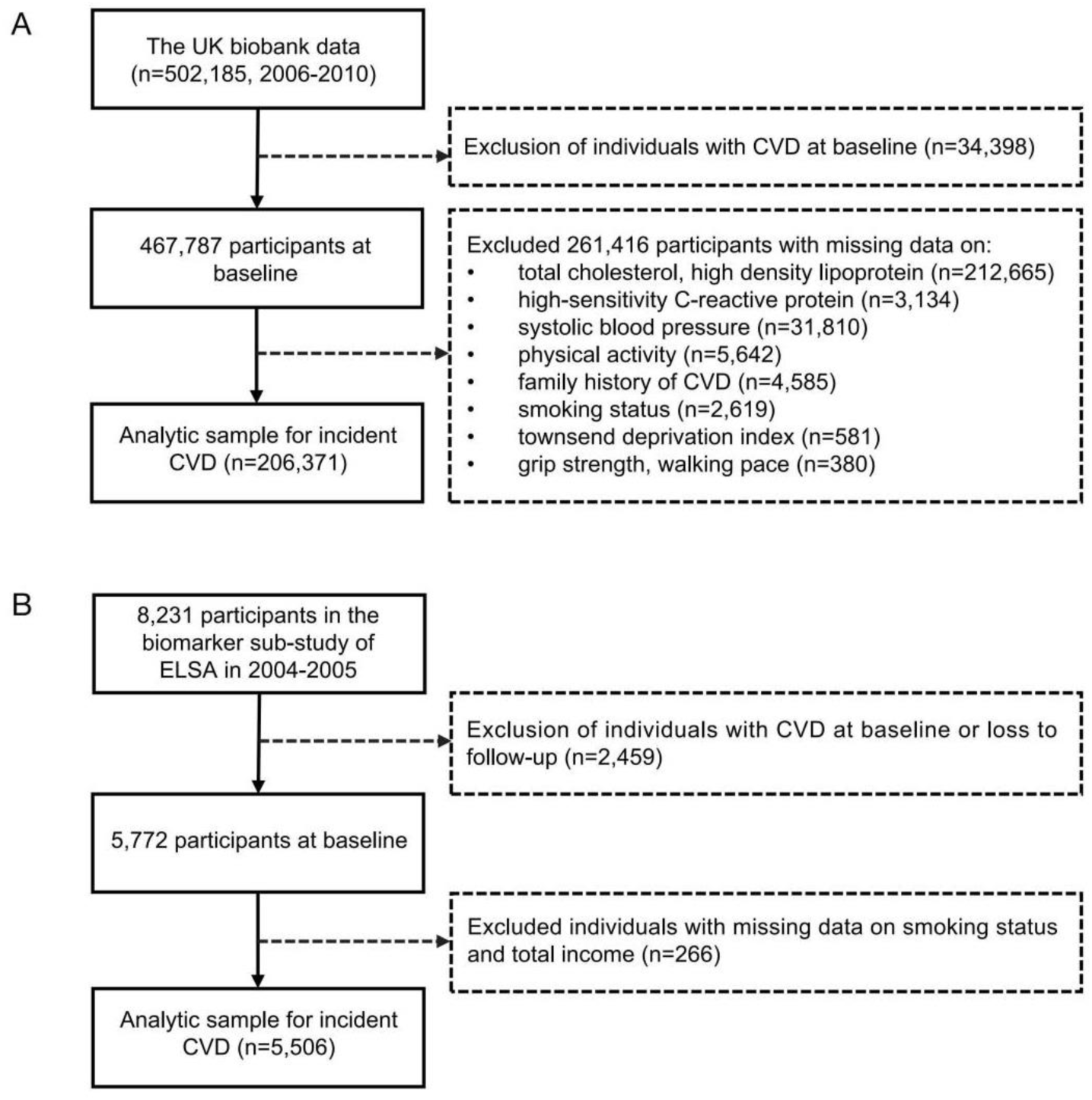
Flowchart of the study population for UKB (A) and ELSA (B). CVD, cardiovascular disease. UKB, UK biobank. ELSA, English Longitudinal Study of Ageing.

The English Longitudinal Study of Ageing (ELSA) was a prospective cohort study conducted in the UK. Details of ELSA methodology and objectives have been reported in the published document[17]. For these participants, face-to-face interviews combined with self-completion questionnaires were conducted by the trained staff to collect data on sociodemographic information, lifestyles, and health-related information. Thereafter, members were surveyed every 2 years using computer-assisted personal interviews and self-completion questionnaires. Furthermore, additional nurse visits were conducted for the assessments of anthropometric (including blood pressure, grip strength, walking pace, and so on) and blood biochemical indicators (including total cholesterol, high-density lipoprotein cholesterol, and so on). We defined wave 2 (2004-2005) as the baseline, because this was the first wave in which core ELSA participants took part in a nurse visit, including 8,231 participants in the biomarker sub-study of ELSA, when biomarkers pertinent to CVD risk were measured. Wave 9 (2018-2019) was the last wave for which data on CVD events were available at the time of our analyses. We excluded participants diagnosed with a CVD event prior to wave 2 (n=1,898) and those without available records of smoking status and income variables (n=827). In total, 5,506 individuals who were free from CVD at baseline and had complete information on all variables were eligible for external validation. Details of the selection process are shown in **Figure 1 (B)**.

### Grip Strength and Walking Pace

Grip strength was assessed in the UKB by a hydraulic hand dynamometer using a Jamar 100105. The isometric grip strength was assessed from single 3-s maximal effort squeeze at the handle of the dynamometer. Both left- and right-hand strengths were measured, and the average value was taken by the sum divided by two. Grip strength was measured in the ELSA by taking 3 measures of both the dominant and nondominant hands of the participants. Grip strength was measured as the isometric handgrip strength in kilograms using a Smedley dynamometer. We used the maximum recorded grip strength from the dominant hand. In the present analysis, we regarded grip strength as a categorical variable according to European consensus on cut-off points for low grip strength (<27 kg for men, <16 kg for women)[18]. Thus, the grip strength was analyzed as a continuous and binary variable in separate analyses.

Walking pace was self-reported using a touchscreen questionnaire completed at the baseline of UKB. The participants who indicated they were able to walk were asked, “How would you describe your usual walking pace?” and offered to choose one of the following: (a) Slow pace; (b) Average/Steady pace; (c) Brisk pace. We defined walking paces from UKB as follows: “Slow pace” was categorized as slow pace, while both “Average/Steady pace” and “Brisk pace” were classified as “normal pace”. In ELSA, participants were asked to walk 8 feet from a standing start on even ground at their usual pace, and the time taken was recorded. The average time of two walks was calculated. We regarded walking pace from ELSA as a categorical variable according to European consensus on cut-off points for “slow pace” (≦0.8 m/s).

### Outcome Ascertainment and Follow-up

The primary outcome for this study was incident CVD. In the UKB, the prevalent CVD was assessed by linked records and self-reported history[19]. The incident CVD was ascertained through linked hospital data until Aug 29, 2021. We used ICD-9 (CVD: 410, 411, 412, 413, 414, 429.79, 430, 431, 432, 433, 434, 435, 436, 437, and 438) and ICD-10 (CVD: I20, I21, I22, I23, I24.1, I25, I46, I60, I61, I63, and I64) to define CVD[20]. In the ELSA, CVD was ascertained based on self-reported physician-diagnosed heart diseases (including angina, heart attack, congestive heart failure, and other heart problems) or stroke.

Follow-up for participants from UKB was initiated from the date of recruitment (between 2006 and 2010), and the follow-up for participants from ELSA was initiated from wave 2 (between 2004 and 2005). The endpoint of follow-up was at the first occurrence of CVD, death, or the censoring date, whichever came first. The censoring date was the date of the last survey in which each participant attended. Ideally, the date of last survey for UKB was Dec 31, 2022, and the last survey for ELSA was wave 9 (2018–2019). Death data were only available up to the wave 6 (2012–2013) in the ELSA. Median and maximum follow-up times of UKB were 13.6 and 15.9 years; median and maximum follow-up times of ELSA were 9.8 and 14.9 years.

### Conventional Predictive Models

The Framingham CVD risk profile[3] estimated 10-year risk of developing CVD events (coronary heart disease, stroke, peripheral artery disease, or heart failure) using key risk factors identified through the Framingham Heart Study. The Reynolds Risk Score[4] was developed for global cardiovascular risk prediction by Reynolds center for cardiovascular research and the center for cardiovascular disease prevention. The ASSIGN score[5] from the Scottish Heart Health Extended Cohort (SHHEC) improved equity in CVD prevention by including social deprivation and family history. The Pooled Cohort Equations (PCEs)[6] was originally designed to calculate 10-year atherosclerotic cardiovascular disease (ASCVD) risk and became the standard for clinical risk assessment for primary prevention of ASCVD[21]. The modifiable variables in the PCEs are also shared risk factors for CVD[22]. Details on the components of the above four conventional predictive models in our analyses are shown in **Supplementary material table 1**.

### Conventional Predictive Models with Recalibration

Due to the different prevalence levels of CVD and the distribution of risk factors in different regions, predictive models often need to be recalibrated to better apply to other groups outside the modeled population[23]. To facilitate a fair comparison between existing conventional predictive models, we recalibrated the four predictive models using the data from UKB. Recalibration was based on calculating the linear predictor for each participant (regression coefficient for each variable × risk factor level) and estimating the baseline survival (10-year survival at-risk factor means) by equating the average 10-year risk to the corresponding observed 10-year Kaplan-Meier rate[24].

### External Validation in ELSA

Added predictive value of grip strength and walking pace in the internal UKB validation data set was externally validated in an independent dataset from ELSA. We categorized selected participants from ELSA into subgroups based on age (Old and Young), grip strength (Normal and Low), and walking pace (Normal and Slow). We transported and applied the optimal model, i.e., the ASSIGN model incorporating grip strength or walking pace to the validation samples from ELSA, applying the same inclusion and exclusion criteria. The robustness of the added predictive value of grip strength and walking pace for predicting CVD was assessed across vulnerable subpopulations. The external validity of the predictions was quantified by performance related to discrimination and reclassification.

### Statistical Analysis

For descriptive statistics, continuous variables were expressed as mean [standard deviation (SD)] or median [interquartile range (IQR)], while categorical variables were expressed as number (percentage). The characteristics of incident CVD were compared using χ^2^ and Kruskal-Wallis tests for continuous and categorical variables, respectively.

To analyze the associations of grip strength and walking pace with incident CVD, Cox proportional hazard regression was used. The hazard ratio (HR) and its 95% confidence interval (95% CI) were calculated from two models: Model 1 was adjusted for age and sex; Model 2 was further adjusted for ethnicity, education level, smoking status, physical activity, BMI, TDI, and prevalent diabetes at baseline based on Model 1.

C statistic and net reclassification improvement (NRI) were used to quantify the model discrimination and added predictive value of grip strength and walking pace beyond that from the conventional predictive models described above. 95% CIs for the C statistic were calculated using the standard error, and 95% CIs for NRI were computed by bootstrapping 500 times the internal-external cross-validation. Sensitivity analyses were conducted after stratifying by sex.

Calibration slope was assessed by comparing expected vs observed event rates in the samples of interest, with 1 indicating the desired mean calibration; values less than 1 signaling risk overestimation; and values more than 1 risk underestimation. Calibration intercept was used to assess whether the model tended to overestimate or underestimate risk. The expected value of calibration intercept was 0; if it was close to 0, the average predicted risks of models were consistent with observed event rates; if it was negative, the model tended to overestimate the risk; if it was positive, the model tended to underestimate the risk.

The missing data on variables from ELSA were imputed using multiple imputations with chained equation. In the ELSA study, the missing rates for walking pace were 2,600 out of 6,333, which is less than 80% of the rate recommended by previous studies[25, 26]. Grip strength, walking pace, SBP, total cholesterol, and HDL were imputed using one imputation model which included age, sex, education, smoking status, drinking status, physical activity, body mass index, C-reactive protein, and anti-hypertension. We performed 10 imputations and generated 10 imputed datasets using the R package “mice”. Effect estimates were computed separately for each of the 10 datasets, and then combined according to Rubin’s rules[25].

We used a two-sided significance level of 0.05 for statistical testing. All statistical analyses were performed using R version 4.3.1 (2023-06-16).

## Results

### Baseline Characteristics of the Study Population

A total of 206,371 participants from UKB (female: 55.6%, median age: 58.0 years [IQR, 50.4-63.4 years]) and 5,506 from ELSA (female: 55.4%, median age: 63.0 years [IQR, 57.0-71.0 years]) were included. During the follow-up, 9.5% (19,664/206,371) of the participants developed CVD in UKB, and 23.1% (1,270/5,506) of participants developed CVD in ELSA. Baseline characteristics of these participants are presented in Table 1. In both the UKB and ELSA studies, participants with incident CVD were older, more likely to use antihypertensive medication, had slow walking pace and exhibited higher levels of TDI, income, and systolic blood pressure compared to those without CVD.

**Table 1.**
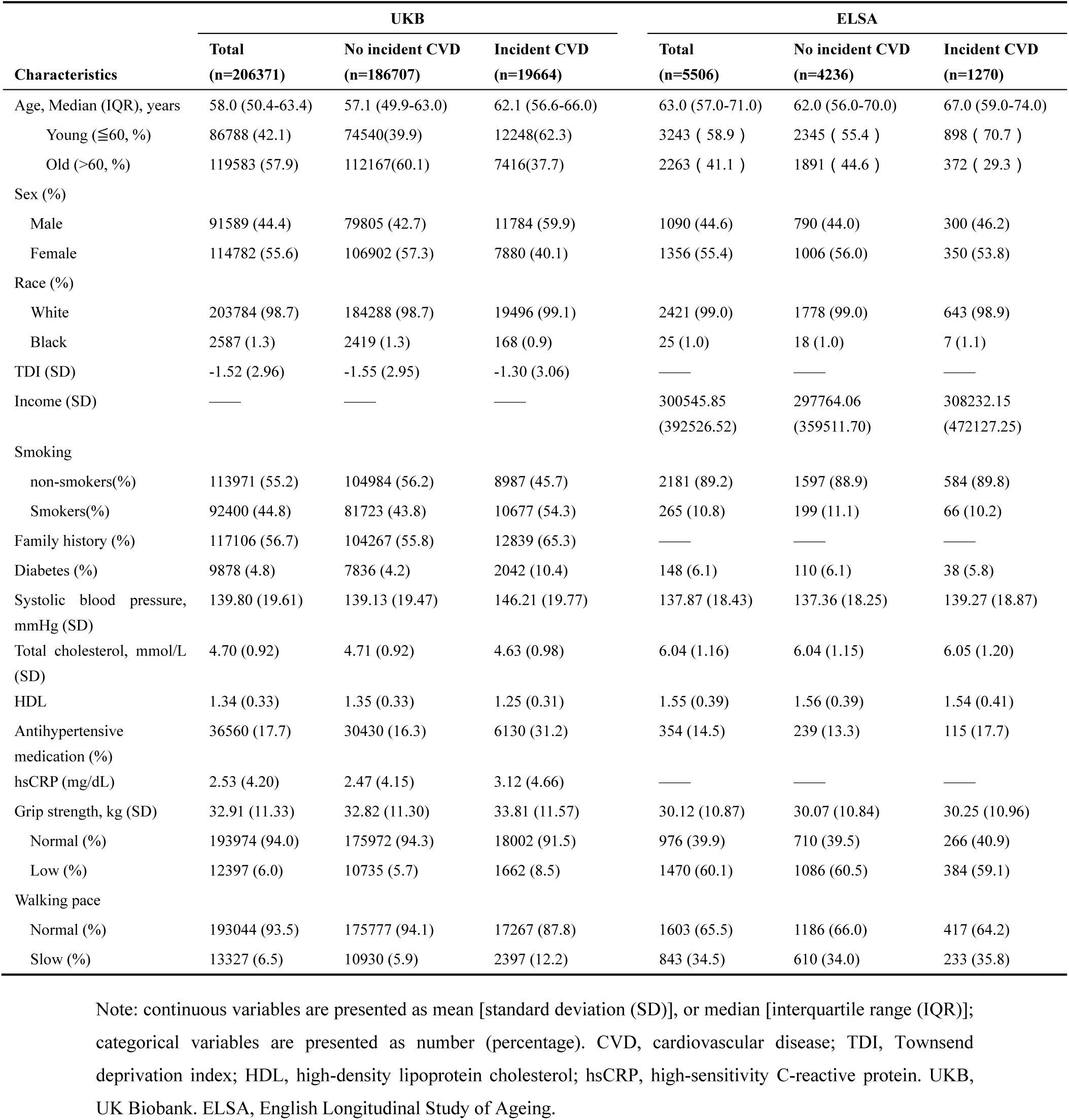
Baseline Characteristics of the Study Population in UKB and ELSA.

### Association of Grip Strength and Walking Pace with Incident CVD

**Figure 2 and Supplementary material table 2** showed the associations of grip strength and walking pace with risks of incident CVD. After adjusting for multiple confounders, participants with low grip strength or slow walking pace had significantly elevated risks of incident CVD than corresponding robust participants (Model 2: low grip strength, HR 1.405, 95% CI 1.338-1.475; slow walking pace, HR 1.844, 95% CI 1.768-1.924).

**Figure 2.**
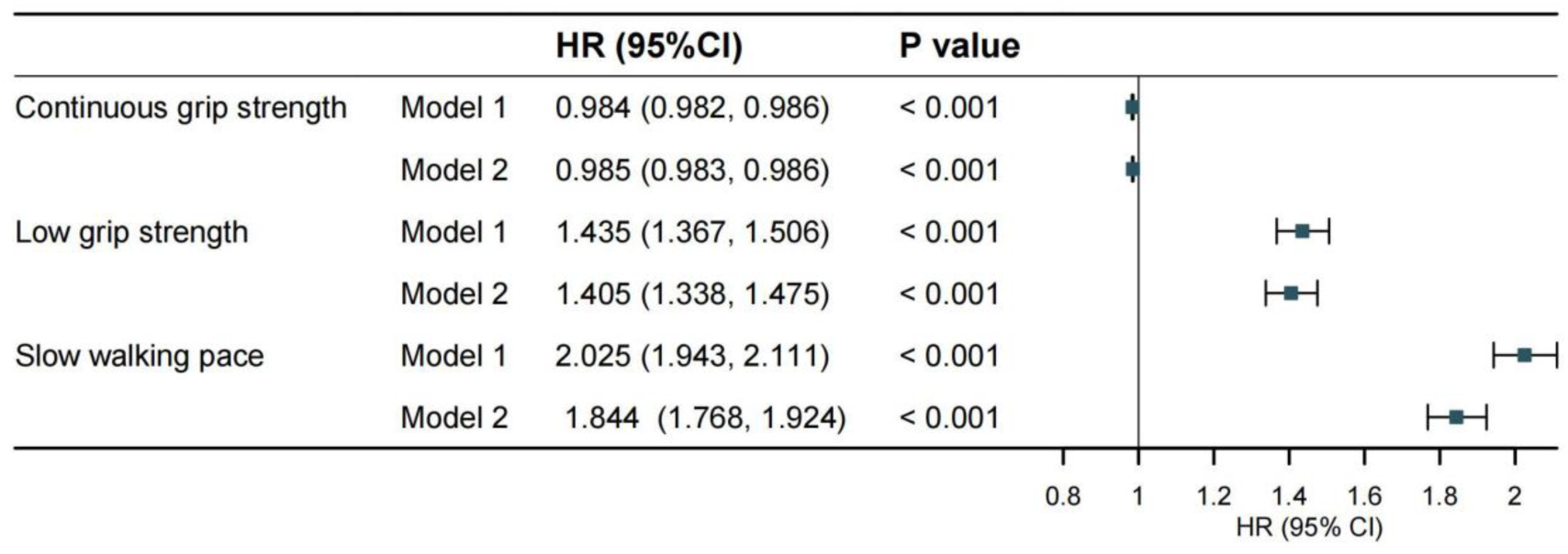
The association of grip strength and walking pace with incident CVD in UKB. Model 1 was adjusted for age and sex. Model 2 was adjusted for age, sex, race, smoking status, drinking status, systolic blood pressure, Townsend deprivation index, and family history of CVD. HR, hazard ratio. CI, confidence intervals.

### Performance of Conventional CVD Risk Prediction Models

In total population, the C index of the four conventional CVD risk prediction models ranged from 0.673 to 0.700. The ASSIGN model performed best with C-index (0.700 [95% CI, 0.693-0.707]) (**Figure 3**). All models demonstrated inferior performance among vulnerable subpopulations compared to robust subpopulations. C index for the ASSIGN model was 0.659 (95% CI, 0.646-0.672) in low grip strength subpopulation vs 0.702 (95% CI, 0.699-0.706) in normal grip strength subpopulation, 0.646 (95% CI, 0.635-0.657) in slow walking pace subpopulation vs 0.701 (95% CI, 0.698-0.705) in normal walking pace subpopulation, and 0.624 (95% CI, 0.614-0.634) in old subpopulation vs 0.701 (95% CI, 0.689-0.712) in the young subpopulation (**Figure 3 and Supplementary material table 3**). Model performance was the worst for old participants with slow walking pace, with C indexes ranging from 0.551 and 0.620 (**Supplementary material table 3**). To account for the possible influence of sex on the results, we repeated this analysis for the males and females separately. The ability to risk prediction was still significantly weaker for participants with low grip strength or slow walking pace, regardless of sex (**Supplementary material table 3**).

**Figure 3.**
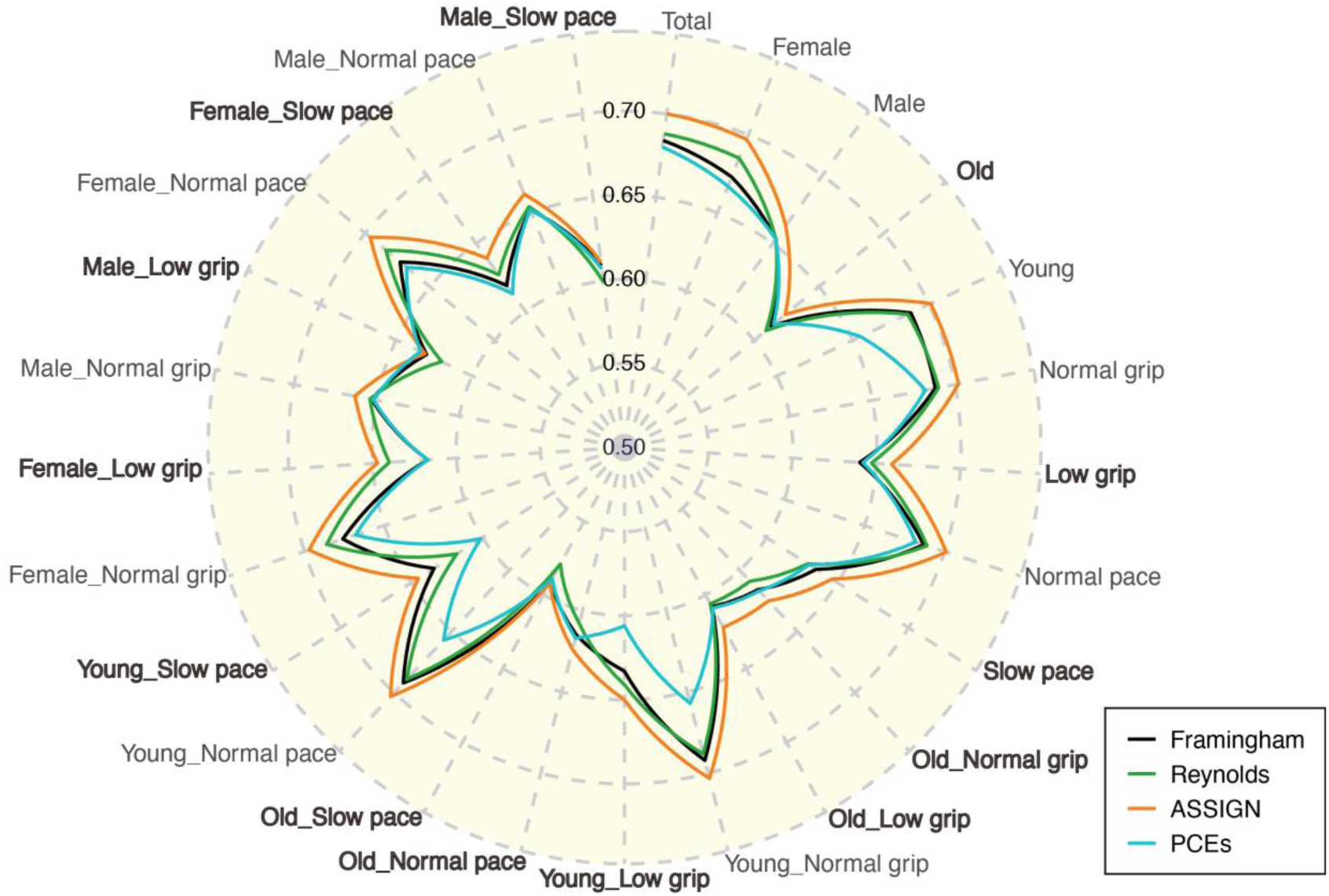
Comparison of C index from models predicting incident CVD according to sex, age, grip strength, and walking pace. The radar plot depicts C indexes for the four conventional CVD risk prediction models (Framingham, Reynolds, ASSIGN, and pooled cohort equations [PCEs]) in total population and 24 subpopulations. Each axis in the radar plot represents total population or 1 subpopulation. Each dashed concentric circle corresponds to a specific value of C index. C indexes from different subpopulations are plotted along each axis and connected to form a polygonal circular curve. Bold represents subpopulations with poor performance of discrimination. Low grip, subpopulation with low grip strength; Slow pace, subpopulation with slow walking pace; Old_Low grip, old population with low grip strength.

The ASSIGN model showed optimal performance with calibration slope of 0.713 (95% CI, 0.685-0.741) and calibration intercept of 0.007 (95% CI, 0.004-0.01) in total population. In contrast, the other three models overestimated the observed risk by large margins: the calibration slopes for Framingham, Reynolds, and PCEs were 0.577 (95% CI, 0.549-0.605), 0.324 (95% CI, 0.291-0.357), and 0.326 (95% CI, 0.308-0.344), respectively; the calibration intercepts were 0.02 (95% CI, 0.017-0.023), 0.031 (95% CI, 0.025-0.036), and 0.028 (95% CI, 0.025-0.031), respectively (**Figure 4A**).

**Figure 4.**
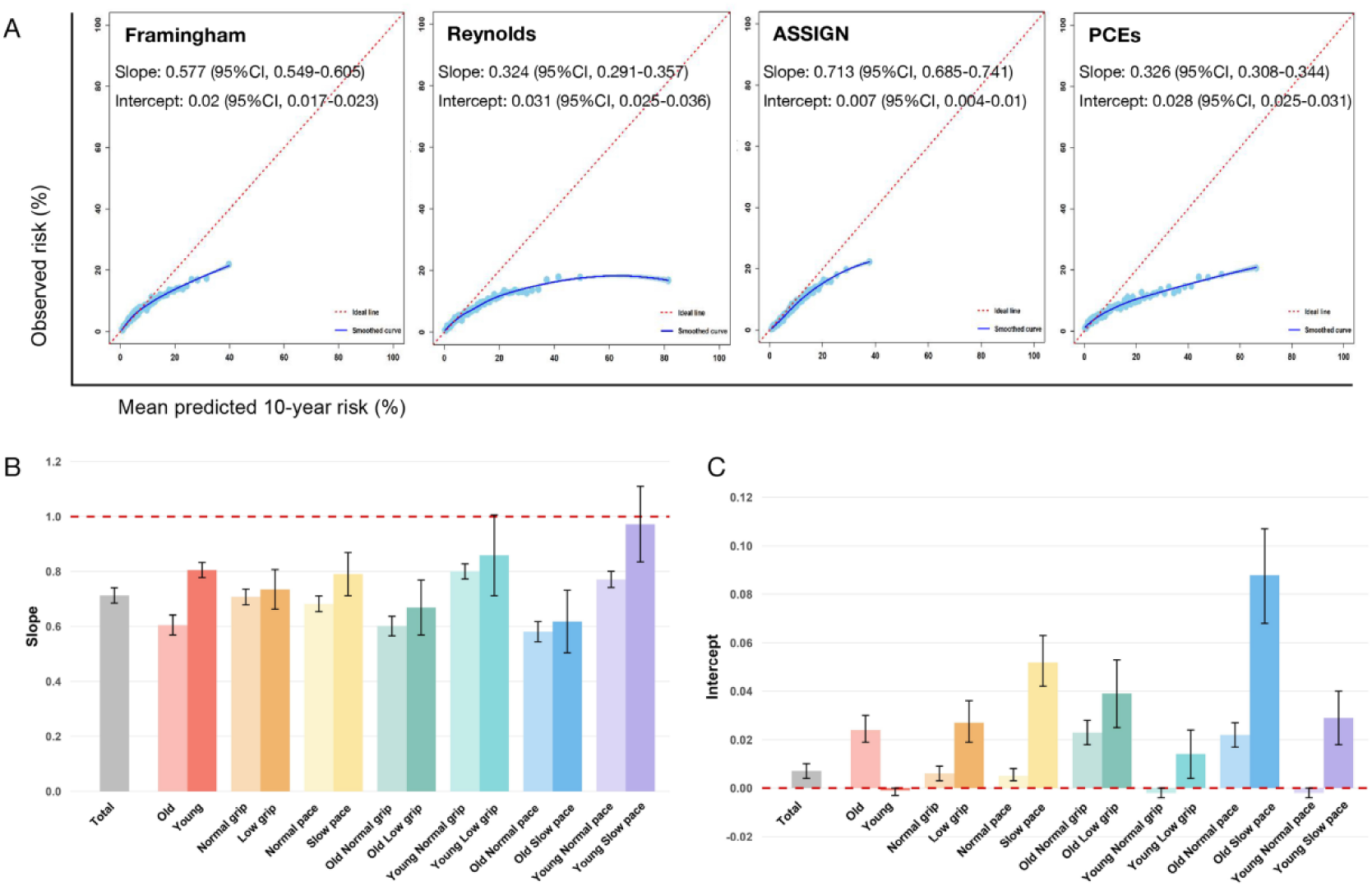
The calibrations of risk prediction models in UKB. (A) Calibration plots for observed vs. predicted risk of CVD as estimated using the Framingham, Reynolds, ASSIGN and PCEs model in overall population, respectively. The dashed line reflects the perfect agreement between observed and predicted risk. (B) Mean values and 95% confidence intervals (95% CI) of calibrations slopes of the ASSIGN model. (C) Mean values and 95% CIs of calibrations intercepts of the ASSIGN model. Error bars on the bar chart represent the 95% CIs. CI, confidence intervals.

Exemplified by the ASSIGN model, there were the significant differences in calibration slope and intercept between the young subpopulation (0.805 [95% CI, 0.778–0.833] and −0.001 [95% CI, −0.003–0]) and old subpopulation (0.605 [95% CI, 0.569–0.641] and 0.024 [95% CI, 0.019–0.03]) (**Figure 4B**). Although calibration slopes in the “Young slow pace” subpopulation were closer to 1 than “Young normal pace” subpopulation (**Figure 4B**), the C index was lower in “Young slow pace” subpopulation (**Figure 3**). And calibration intercepts were closer to 0 within subpopulations with normal grip strength and walking pace (**Figure 4C**), indicating that the systematic bias of the ASSIGN model in these subpopulations is smaller than that within vulnerable subpopulations. The trend in differential calibration performance across above subpopulations was similar for the Framingham, Reynolds, and PCEs models (**Supplementary material table 4 and 5**). Overall, the calibration plots displayed poor predictive accuracy for most risk models in old subpopulation or vulnerable subpopulations.

### Added predictive value of grip strength and walking pace

The addition of grip strength and walking pace to conventional risk prediction models was associated with an increase in the C statistics (C index change) for CVD events (**Supplementary material table 6)**. Especially, the addition of both continuous grip strength and walking pace to the ASSIGN model (i.e., ASSIGN plus grip_walking pace) ameliorated discrimination in vulnerable subpopulations, e.g., C index change was 0.009 (95% CI, 0-0.017) for old subpopulation and 0.009 (95% CI, 0.001-0.018) for old subpopulation with low grip strength in training set (**Figure 5A**). Although the C index change exhibited only modest increases in old subpopulations and old subpopulations with low grip strength in the testing set, the changing trend was similar to the training set (**Figure 5B**). The C statistic was blamed as being insensitive[27, 28], thus, we further assessed the added predictive value of grip strength and walking pace using the reclassification analyses.

**Figure 5.**
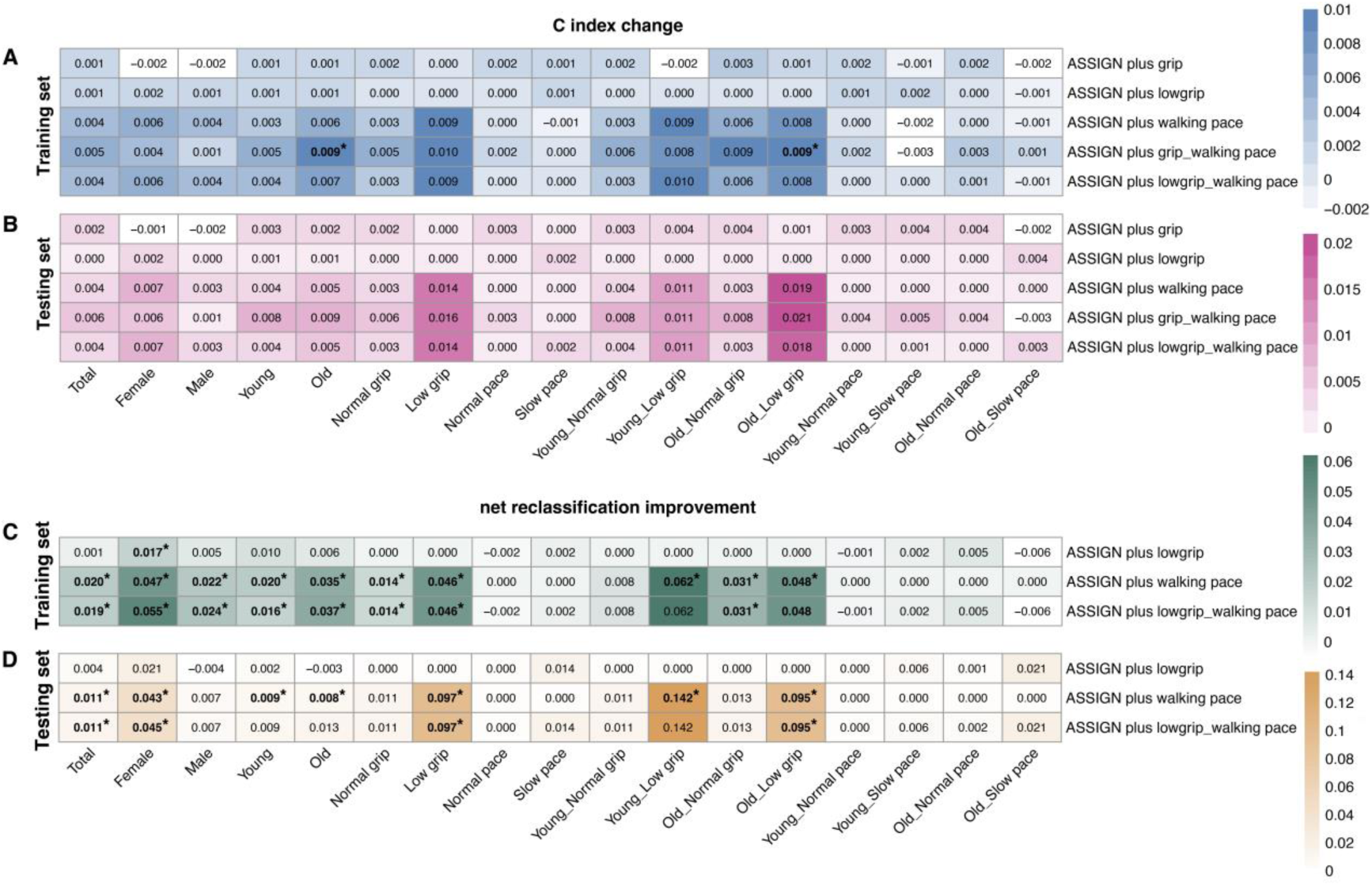
Heat maps displaying the changes in the C index changes and NRIs after adding grip strength and walking pace to the ASSIGN model in UKB. The values of C index changes and net reclassification improvements across different subpopulations were scaled, with darker colors indicating larger values. (A) C index change in the training set, (B) C index change in the testing set, (C) NRI in the training set, (D) NRI in the testing set; n=144,460 in total training set, n=61,911 in total testing set. \**P* < 0.05. NRI, net reclassification improvement. UKB, UK Biobank.

Adding walking pace to the ASSIGN model showed marked positive NRI values for vulnerable subpopulations, e.g., old subpopulation, subpopulation with low grip strength, and their combination (**Figure 5C-D**). Notably, the NRI for the walking pace was 0.097 (95% CI, 0.005-0.177) in subpopulation with low grip strength (**Figure 5D and Supplementary material table 7**). Unexpectedly, there was hardly any improvement in the subpopulation with slow walking pace adding grip strength to the conventional prediction model for either C index change (**Figure 5A-B**) or NRI (**Figure 5C-D**) analysis.

### External validation among vulnerable subpopulations in ELSA

We further investigated whether the observed findings remained robust in an independent external sample, considering that the majority of participants were of White and England descent in both UKB and ELSA. Our analysis revealed a decline in the predictive accuracy of the ASSIGN model for CVD across all subpopulations in ELSA, potentially related to a more advanced age profile and the lack of certain variables (e.g., family history) in the ELSA. Notably, similar to the findings from UKB, the inclusion of walking pace demonstrated a trend towards an increased predictive value for old subpopulation (C index change, 0.004 [95% CI, −0.022-0.03]; NRI, 0.001 [95% CI, −0.061-0.07]) and the subpopulation with low grip strength (C index change, 0.002 [95% CI, −0.027-0.031]; NRI, 0.042 [95% CI, −0.056-0.09]), although this did not achieve statistical significance (**Figure 6, Supplementary material table 8 and 9**). We propose that considerable differences in the age distribution (UKB: median, 58.0 years [IQR, 50.4-63.4 years]; ELSA: median, 63.0 years [IQR, 57.0-71.0 years]) and 10-year CVD incidence rate (UKB: 9.5 cases per 1000 person-years; ELSA: 23.1 cases per 1000 person-years)] may play the significant roles. Besides, the absence of the family history variable in ELSA, the different definitions of walking pace between UKB and ELSA, and the substitution of the income variable from ELSA for the Townsend Deprivation Index (TDI) variable from UKB may contribute to the discrepancies observed in the results between ELSA with UKB.

**Figure 6.**
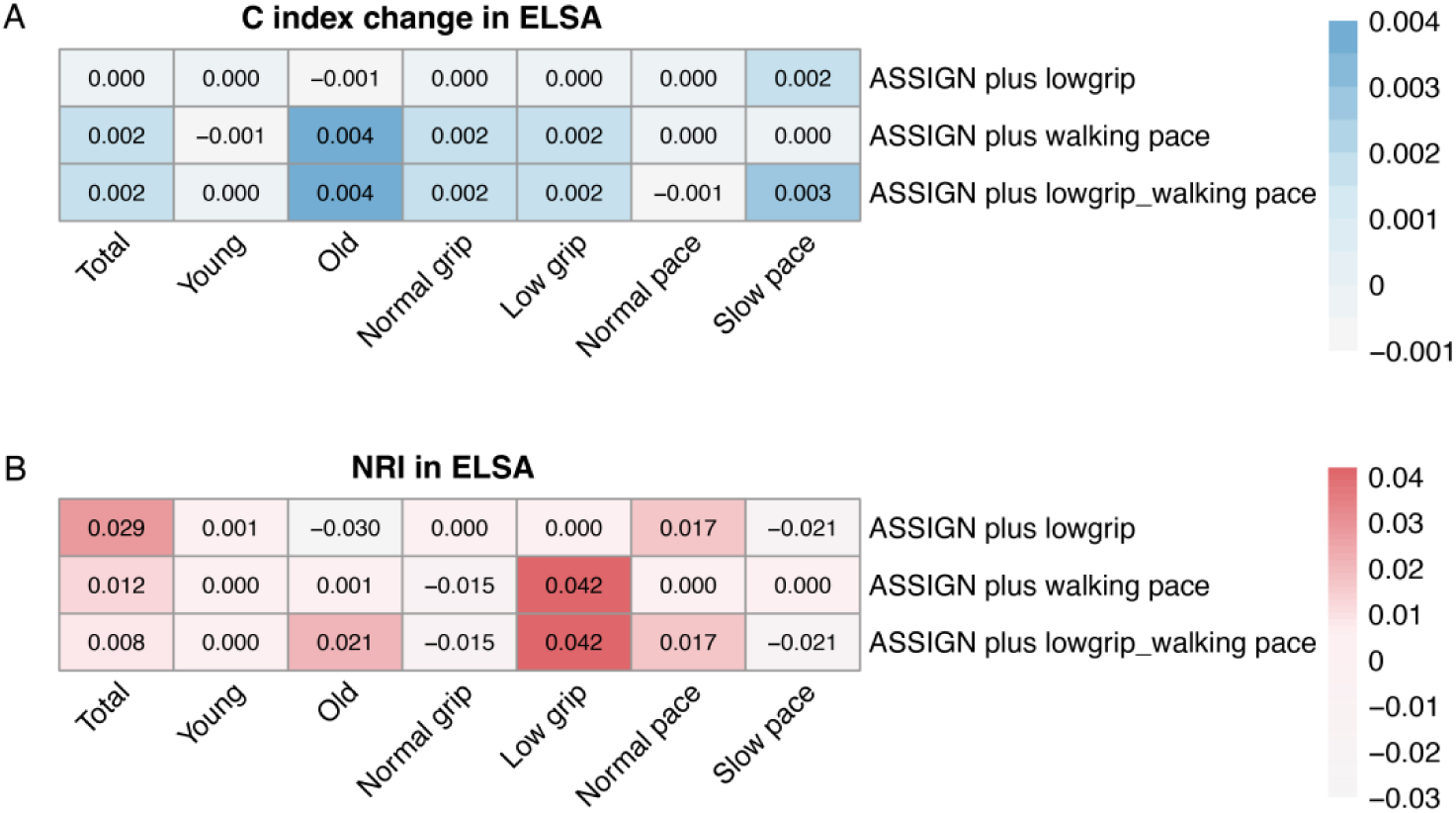
Heat maps displaying the changes in the C index changes and NRIs after adding grip strength and walking pace to the ASSIGN model in ELSA. The values of C index changes and NRIs across different subpopulations were scaled, with darker colors indicating larger values. (A) C index change; (B) NRI in training set, n=5506 in total. NRIs, net reclassification improvements. NRI, net reclassification improvement. ELSA, English Longitudinal Study of Ageing.

## Discussion

In a large-scale cohort from the UK Biobank, we evaluated the performance of four conventional CVD risk prediction models (Framingham, Reynolds, ASSIGN, and PCEs) and had the following findings: all four models did not work well in vulnerable subpopulations with advanced age, low grip strength, or slow walking pace, with the ASSIGN model demonstrating relatively better performance; adding walking pace as a variable into the ASSIGN model significantly enhanced its predictive capability in vulnerable populations, which was further external validated in the ELSA. This study underscores the effectiveness of walking pace as a simple and easy-to-implement tool for CVD risk prediction among vulnerable subpopulations.

Advanced age, low grip strength, and slow walking pace represent vulnerable status as routine availability in cohort studies and are also identified as strong risk factors in earlier prediction of multiple age-related diseases[29–31]. Consistent with previous studies[14],[32], the associations of grip strength/walking pace and the risk of incident CVD were observed, supporting the viewpoint on elevated risks of incident CVD in participants with low grip or slow pace as compared with robust participants. Prior studies showed that the effect of conventional risk factors is attenuated with increasing age[7], and the application of conventional predictive models has been prevalent in this research domain, providing stratified demographic and epidemiological data[7, 33], e.g., race, age, and sex, yet frequently neglecting the critical physical function heterogeneity that is essential for a comprehensive understanding. In this study, we defined vulnerable subpopulations characterized by advanced age, low grip strength, and slow walking pace to compare the performance of conventional CVD models among those subpopulations, thereby effectively filling a gap in this area.

Our study revealed that adding grip strength and walking pace into the conventional prediction model provided discrepant incremental predictive values among some vulnerable subpopulations. The inclusion of walking pace to conventional risk factors led to the optimal improvement for incident CVD among subpopulations with low grip strength. Comparable results from ELSA suggested the added predictive value of walking pace remains robust within vulnerable subpopulations for CVD risk. Moreover, cost-effectiveness should eventually guide us in considering that walking pace, a simple measurement, may be necessary enough to improve predictive performance in clinical practice[34].

### Clinical and public health implications

These data indicate that walking pace could have a useful role in CVD risk prediction. Walking pace is a clinically inexpensive, rapid, non-invasive, and accessible indicator, which makes the measurement of walking pace easily implementable in various application settings, such as community centers, hospitals, and nursing homes. This also enables more accurate identification of high-risk individuals, facilitates early intervention, and allows for the personalization of care plans, ultimately improving CVD health outcomes in vulnerable groups. Further research should replicate these findings in other subpopulations from more cohorts and application settings, and both real-world data and clinical trials are needed to explore the optimal combination of conventional risk predictors, grip strength, and walking pace, as well as the most effective interventions for vulnerable subpopulations in CVD prevention practice.

### Limitations

Several limitations should be taken into account. First, the ascertainment of CVD in the ELSA was based on self-reported physician-diagnosed information, which might lead to a misclassification bias. However, Xie et al. confirmed that 77.5% of self-reported coronary heart disease were consistent with the medical records in the ELSA[35]. This suggested that the potential misclassification bias was minor. Second, although we adjusted for multiple confounders, residual confounding might remain, such as genetic susceptibility and diet. In addition, the probability of type I error might be inflated, because several analyses conducted multiple comparisons without any adjustment for the multiplicity. However, we performed sensitivity analyses stratifying by sex, and above results were consistent with the main analyses (**Supplementary material table 3-9**). Finally, although our study encompasses a broad range of locations within Europe, the global generalizability and internal validity of our findings need further validation in other cohort studies and settings. Future research efforts should verify and expand on our results in various locations and different contexts.

## Conclusions

Low grip strength and slow walking pace are associated with an increased risk of incident CVD, with walking pace showing a stronger association. The performance of the conventional CVD risk prediction models was worse in vulnerable subpopulations characterized by advanced age, low grip strength, and slow walking pace. Walking pace optimized the performance of the conventional risk prediction model among vulnerable subpopulations. These findings suggest walking pace, a low-cost and non-invasive indicator, could be assessed to improve the prediction of incident CVD among vulnerable subpopulations with advanced age or low grip strength in community settings. By providing nursing practice with a practical tool to identify high-risk individuals, this study enables personalized care plans and early intervention and ultimately supports equitable health policies.

## Abbreviations

UKB: UK Biobank
CVD: Cardiovascular disease
ELSA: English Longitudinal Study of Ageing
PCEs: Pooled Cohort Equations
C index: Harrell’s concordance index
NRI: Net Reclassification Improvement
CVD: Cardiovascular Disease
IHD: Ischemic Heart Disease
IS: Ischemic Stroke
PAD: Peripheral Artery Disease
SD: standard deviation
TDI: Townsend deprivation index
HDL: high-density lipoprotein cholesterol
hsCRP: high-sensitivity C-reactive protein.

## Acknowledgements

This research has been conducted using the UK Biobank Resource under application number 61856. All authors thank the original data collectors, depositors, copyright holders, and funders of UK Biobank and English Longitudinal Study of Ageing.

## Author contributions

WL performed formal analysis, writing–original draft preparation, visualization, methodology, and funding acquisition. LX contributed to formal analysis, methodology, and data curation; JX performe writing–review & editing and data Curation. ZY performed visualization and methodology. SJ and LZ revised it critically for important intellectual content and performed funding acquisition. All authors read and approved the final version of the manuscript.

## Data availability

The UK Biobank data that support the findings of this study can be accessed by researchers on application (www.ukbiobank.ac.uk/). Data supporting the results of external validation are available from official websites of English Longitudinal Study of Ageing (https://ukdataservice.ac.uk).

## Declarations

### Ethics approval and consent to participate

The UK Biobank obtained ethics approval by the Northwest-Multicentre Research Ethical Committee that had obtained written informed consent from all participants. And the English Longitudinal Study of Ageing obtained ethics approval by the National Research and Ethics Committee that had obtained written informed consent from all participants.

### Consent for publication

Not applicable.

